# Rethink the Flow: Establishing a Preanalytical Workflow for Protein Detection in Menstrual Effluent

**DOI:** 10.64898/2026.11.19.26368425

**Authors:** Shekemi Denuga, Barbara Lukaszczyk, Theresa Okwuegbuna, Kezia E. Suryoraharjo, Alana F. Ogata

## Abstract

**BACKGROUND:** Menstrual effluent (ME) is a promising, minimally invasive biofluid that contains unique protein biomarkers indicative of systemic and gynecological health. Despite its potential, the adoption of ME as a biofluid for disease identification remains limited because no standardized protocols exist for sample collection, processing, and storage. The lack of standardization remains a major challenge in ME research, introducing non-biological variability that limits accurate protein measurements and downstream interpretation for disease diagnosis. We examine how preanalytical factors, including centrifugation, freeze-thaw cycling, and pre-processing storage conditions, affect sample stability and protein concentration measurements.

**METHODS:** Single-molecule array (Simoa) technology was used to quantify cancer antigen 125 (CA125) in ME samples collected from 19 participants.

**RESULTS:** We found that delays in sample processing beyond eight hours increased the probability of unsuccessful fractionation (*p* < 0.02), a metric of sample stability. We detected CA125 in 100% of ME and ME-derived plasma samples; however, plasma showed better reproducibility across aliquots. Repeated freeze-thaw cycling significantly altered CA125 concentrations in plasma samples. ME samples stored at 4°C before centrifugation maintained their ability to fractionate and protein integrity for up to 120 hours. We also demonstrate that the developed workflow is compatible with longitudinal monitoring of CA125 in ME over three consecutive cycles.

**CONCLUSIONS:** These results support at-home self-collection of ME for sample acquisition. Furthermore, the developed workflow enabled reliable protein biomarker analysis, facilitating the use of ME as a biofluid for biomarker discovery for gynecological diseases.

## INTRODUCTION

Menstrual effluent (ME), commonly referred to as menstrual blood, is a complex biofluid comprising of blood, cervical mucus, shed endometrial tissue, and secreted factors, including proteins, immune cells, and nucleic acids, that reflect the uterine microenvironment (1). Historically regarded as biological waste, ME has garnered interest since 2007 as a promising biofluid containing biomarkers indicative of systemic and gynecological diseases (2–4). ME has been shown to contain up to 385 unique protein biomarkers not found in venous blood or vaginal fluid proteomes (5). Consequently, researchers are investigating ME to identify biomarkers linked to physiological and pathological conditions of the female reproductive system, including high-risk Human Papillomavirus (HPV) infections and endometriosis (6). Detecting nucleic acids indicative of cervical abnormalities and HPV infections is among the most advanced diagnostic applications of ME, with sensitivity and specificity often exceeding 80% (7). Uncontrolled preanalytical variables, such as the day of sample collection, can affect the efficiency of detecting HPV DNA and cervical intraepithelial neoplasia grade (CIN). Lee et al. demonstrated that, when stratified by CIN grade, sensitivity for detecting CIN 3 or worse was 87.5% on day 1 and 62.5% on day 2 of the menstrual cycle (8). Such preanalytical variability may also explain inconsistent results in protein biomarker discovery studies for diseases like endometriosis. For example, Manan et al. used undisclosed sampling and processing methods and associated elevated vascular endothelial growth factor-A (VEGF-A) concentrations in ME with endometriosis (9). However, Malik et al. found no differences in VEGF-A concentrations in day-two menstrual cup-collected samples from individuals with and without endometriosis (10).

Preanalytical factors, including sample collection method, collection day, sample processing, storage, and transportation, can alter sample properties (e.g., through degradation), influence biomarker measurements, and, consequently, dictate biological interpretation (11,12). Establishing guidelines for preanalytical ME handling will enable reliable biomarker quantification, reproducible biomarker identification, and accurate disease diagnosis (13). However, to the best of our knowledge, no systematic investigation has examined how preanalytical variability affects protein biomarker measurements in ME.

In this work, we developed and optimized a menstrual cup self-sampling protocol for ME. We examined how key preanalytical factors, including freeze-thaw (FT) cycling and storage conditions, affect ME stability and protein analysis. We used CA125 as a model protein biomarker because of its abundance in ME (14) and its association with endometriosis (15), cervical cancer (16), and ovarian cancer (17). We quantified CA125 concentrations in ME samples using single-molecule array (Simoa) technology, achieving a detection limit of 0.1 ± 0.1 pg/mL (*N* = 19). We found that FT cycling increased sample variability, whereas storage at 4 °C extended ME sample stability for up to 120 hours. Finally, we demonstrated the method’s compatibility with longitudinal CA125 monitoring over three consecutive cycles. This work establishes recommendations for standardized guidelines for ME collection and processing, supporting ME as an accessible, minimally invasive biofluid for quantifying and monitoring protein biomarkers for gynecological disease diagnosis.

## METHODS

### ME Sample collection

ME was self-collected by 19 individuals under a protocol approved by the University of Toronto (UofT) Research Ethics Board (Human Protocol #00049006). Eligible participants were individuals aged ≥18 who menstruate and could pick up and return a kit at UofT Mississauga. Individuals under 18 or who did not menstruate were excluded. Participants provided written informed consent and received a menstrual cup kit to collect ME for 3-4 hours on their heaviest flow day (Supplementary Figure 1). Participants transferred the sample to the collection tube, placed it in a biohazard bag and returned it to the laboratory within 24 hours of starting collection.

### Sampling Processing

ME samples were processed within four hours of drop-off. Samples were aliquoted and centrifuged at 2,000 × g for 20 minutes at 4°C to separate the cellular pellet from the acellular supernatant. A minimum of 50 µL of ME was stored as unprocessed ME. After processing, samples were immediately stored at −80°C for at least 24 hours before analyses.

### Simoa Measurements

All measurements were performed using the CA125 assay previously reported on the HD-X Analyzer (Quanterix) according to the manufacturer’s three-step protocol (Supplemental Methods, S1) (18). We used a CA125 assay kit purchased from R&D Systems (cat# DY5609-05). Each independent run included nine levels of recombinant protein standards, ranging from 0.41-900 pg/mL, prepared in Homebrew Detector/Sample Diluent to generate a calibration curve (Supplemental Figure 2, Supplemental Table 4). The measured Average Enzyme per Bead (AEB) values were fit against the calibrator concentrations using a 4-parameter logistic curve weighted by 1/y^2^ to generate calibration curves. The three quality control samples at 540 pg/mL, 54 pg/mL, and 5.4 pg/mL, prepared in Homebrew Detector/Sample Diluent, were also included in each run, with acceptable error margins below 20%. All calibrators and samples were measured in duplicate, with CA125 levels for each sample determined using the calibration curve prepared on the day of analysis.

### Quantification of CA125 in ME and Plasma

Each sample was thawed on ice for at least one hour, vortexed for 30 seconds, and briefly centrifuged to ensure homogeneity. The assay was validated using pooled and individual ME and plasma samples (Supplemental Information, S3). ME samples were diluted 60,000-fold, and plasma samples 8,000-fold in Homebrew Detector/Sample Diluent. To validate the accuracy of CA125 measurements, each sample was analyzed unspiked and after spiking with 2 pg/mL, 10 pg/mL, and 100 pg/mL of recombinant CA125. Percent recovery at each spike level was calculated and averaged across the three spike levels with an acceptance criterion of 70-130%. Samples below the acceptable range were excluded from this study. Total CA125 concentration in the sample was calculated by multiplying the Simoa concentration by the dilution factor.

### FT Cycling

Eight plasma samples were measured for baseline CA125, then subjected to two sequential FT cycles. Each cycle consisted of storage at –80°C for at least 24 hours, followed by thawing on ice, CA125 quantification, and return to – 80°C for the next cycle.

### Varying Storage Conditions

Four ME samples were aliquoted and stored at ∼20°C, 4°C, or −20°C for up to 120 hours. At 6, 24, and 120 hours, one aliquot per sample and condition was centrifuged to assess fractionation. If successful, plasma was aliquoted and CA125 concentration measured.

### Data Analysis

The HD-X analyzer software calculated sample concentrations using the calibration curves generated on the day of the measurement. Data are reported as mean ± standard deviation (SD) of the measurement, over *N* replicates, unless specified otherwise. The limit of detection (LOD) was calculated as the signal at zero analyte concentration plus three SDs of the first calibrator (0.4 pg/mL). All data analysis and figure generation were performed in GraphPad Prism version 11.1.0. The Mann-Whitney test, Spearman correlation analysis, Wilcoxon test, and Bland-Altman analysis were performed in GraphPad Prism to assess statistically significant differences in sample collection variables and CA125 concentrations in ME and plasma. Statistical significance was set at a two-tailed *p-* value < 0.05.

## RESULTS

ME self-sampling kits and protocols were designed using a user-centred approach, including iterative user engagement surveys (Supplemental information S2.1). For the self-sampling of ME, participant demographic characteristics are summarized in Supplemental Information S2.2. The median self-reported heaviest flow day was Day 2 of menstruation, consistent with the general understanding that flow peaks early in the cycle (Supplemental Table 2). The median menstrual cup wear time was 3 hours 24 minutes, within the requested 3-4 hours window. However, 35% of samples (*N* = 9) were collected after the cup was worn longer than requested.

ME samples varied widely in physical properties such as volume, colour, viscosity, and degree of clotting (Figure 1A-B). The volume of ME collected ranged from 0.9 to 7.0 mL, with a median of 2.0 mL, consistent with previous studies (19). We observed no correlation between the volume of ME collected and the length of cup wear (ρ = -0.3, *p* = 0.2) (Figure 1C). Of the 26 ME self-collected samples, 81% (21/26) fractionated into two distinct layers, while the remaining 19% (5/26) failed to fractionate, a metric we can use to approximate sample stability. Samples that failed to fractionate had longer intervals between sample collection and drop-off (*p* = 0.02) (Figure 1D-E).

**Figure 1.**
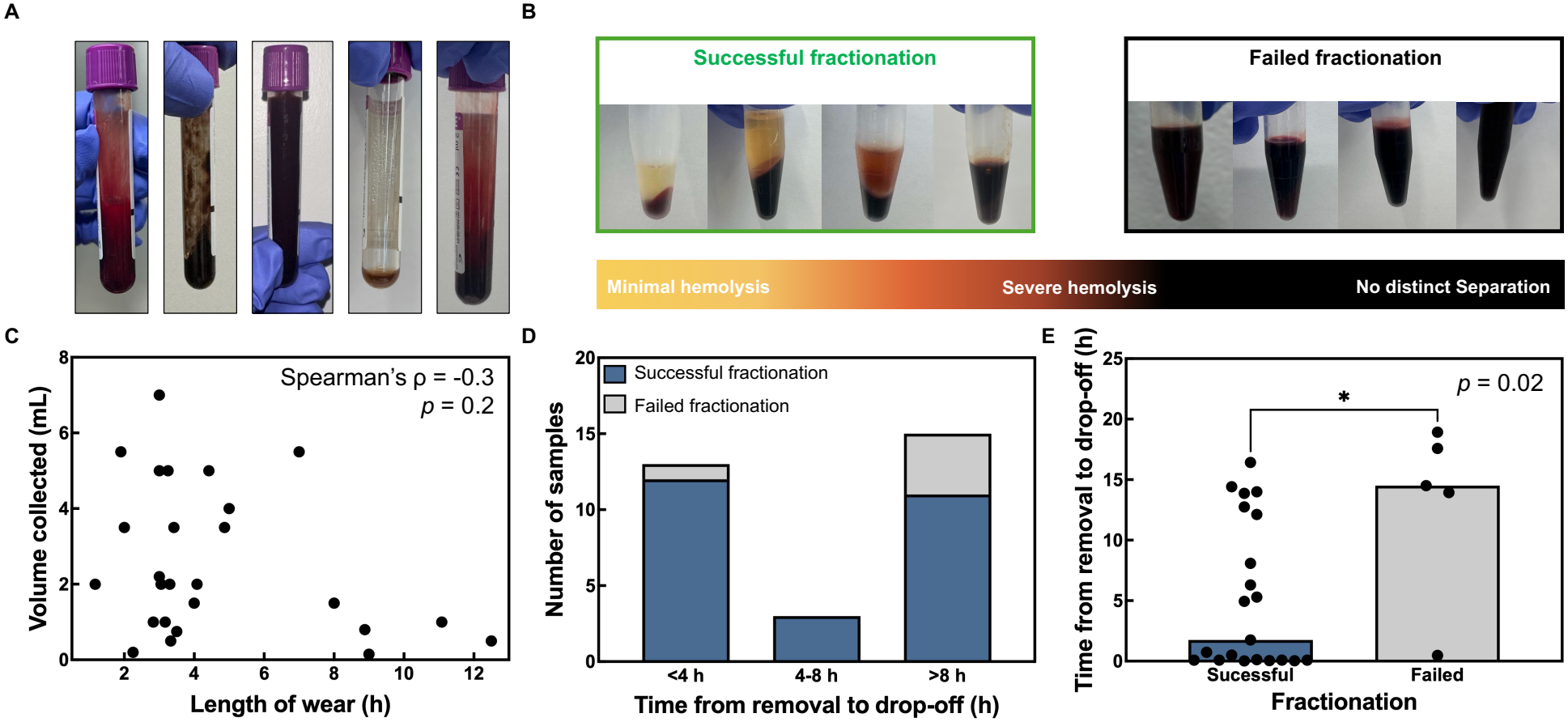
Images of ME samples (A) in collection tubes (B) after centrifugation, with a hemolysis reference. (C) Correlation between ME volume and menstrual cup wear duration. (D) Number of samples stratified by time from cup removal to drop-off. (E) Time from cup removal to sample drop-off, stratified by fractionation outcome (*p* < 0.05). Bar plots indicate the median. Differences in medians were assessed by the Mann-Whitney test.

We developed a Simoa-based protocol to quantify CA125 in ME, compatible with detecting low-abundance protein biomarkers for diagnosing gynecological diseases. The CA125 assay achieved a limit of detection of 0.1 ± 0.1 pg/mL (*N* = 19) with an inter-assay variability of 6% and intra-assay variability of 4% (Figure 2A; Supplemental Table 4-6). As not all samples fractionated successfully, we developed and validated two parallel workflows for protein detection in ME and plasma (Figure 2B).

**Figure 2.**
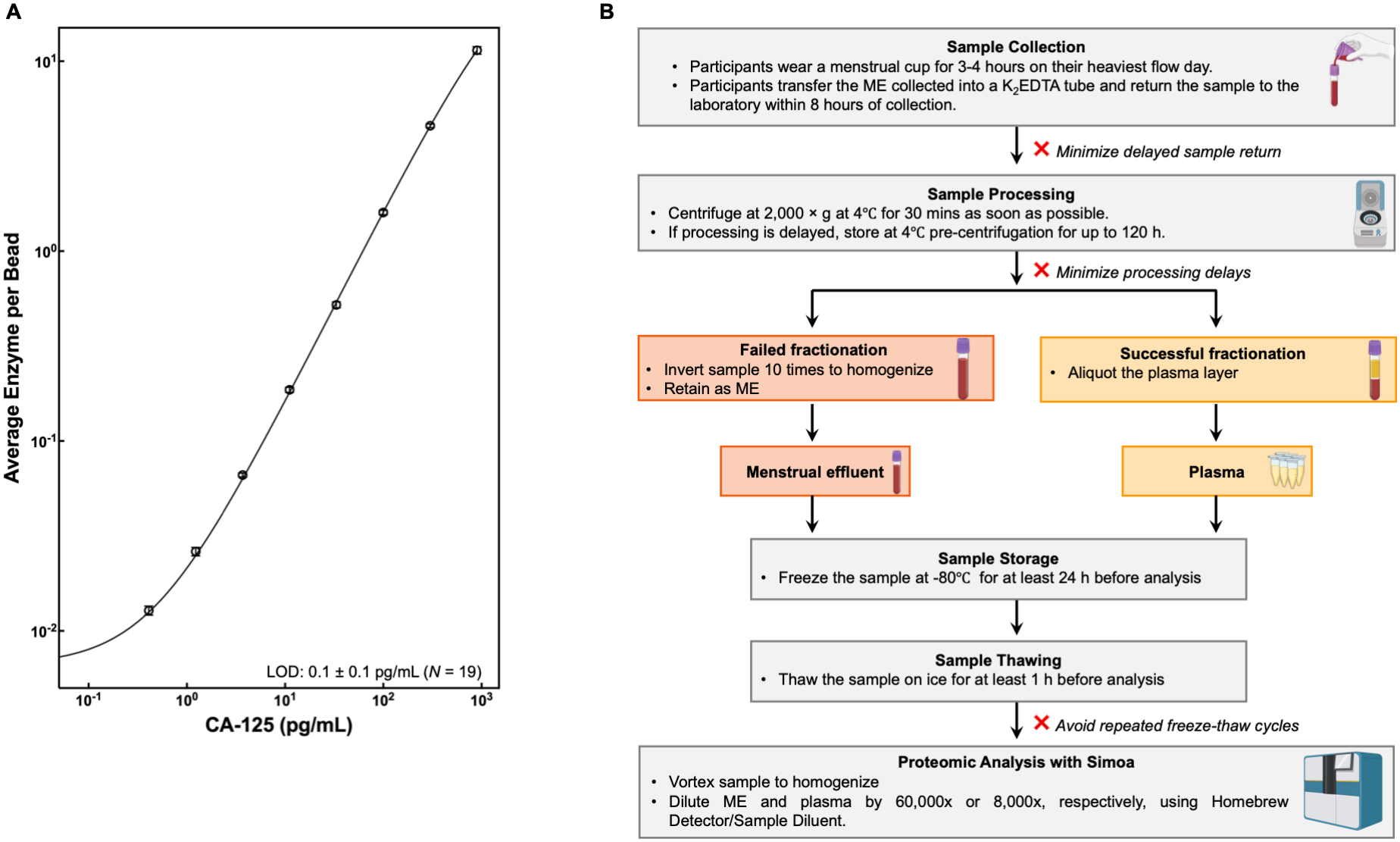
(A) The mean CA125 response calculated from 19 independent Simoa calibration curves. Error bars are the SD at each concentration across 19 replicates. (B) The flowchart of ME sample processing from ME collection to Simoa analysis.

### Plasma Provides Low Aliquot-to-Aliquot Variability

We investigated the reproducibility of CA125 concentrations in ME and plasma samples using two randomly selected aliquots (*N* = 4 ME, *N* = 3 plasma). Each aliquot was vortexed for 30 seconds to ensure homogeneity before CA125 concentrations were measured. The mean coefficient of variation (%CV) for ME aliquots was 30% ± 20% (range: 1–51%), compared with 0.9% ± 0.2% for plasma (range: 0.7–1.2%). The aliquot-to-aliquot variability observed in ME samples exceeds the intra-assay and inter-assay variability of 4% and 6%, respectively (Figure 3, Supplemental Table 8). Based on its low variability, plasma was selected as the focus for the remainder of this work.

**Figure 3.**
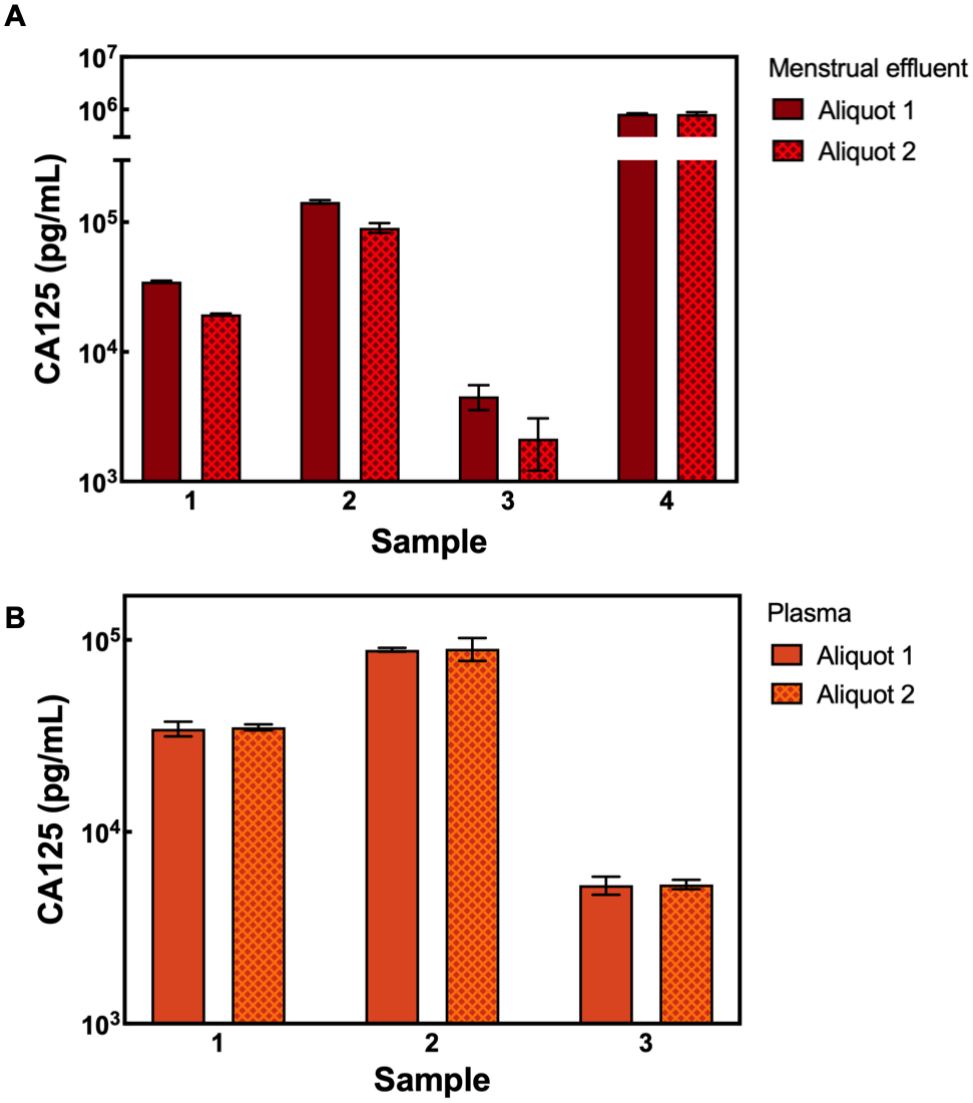
Aliquot-to-aliquot variability of CA125 in (A) ME aliquots (%CV 40%, 32%, 51%, and 1%, respectively) and in (B) plasma samples (%CV 1.2%, 0.9%, and 0.7%, respectively). Each data point is an average of two replicates corrected by the dilution factor, with error bars representing the SD.

### FT Cycling Introduces Variability in Plasma CA125 concentrations

As shown in Figure 4, plasma samples subjected to three consecutive FT cycles showed inconsistent changes in CA125 concentration (Supplemental Table 9). At FT cycle 2, CA125 concentration remained unchanged in one sample, increased in four, and decreased in three of the eight samples analyzed. After FT cycle 3, one sample returned to baseline, while five increased, and two fell below their baseline (Figure 4B).

**Figure 4:**
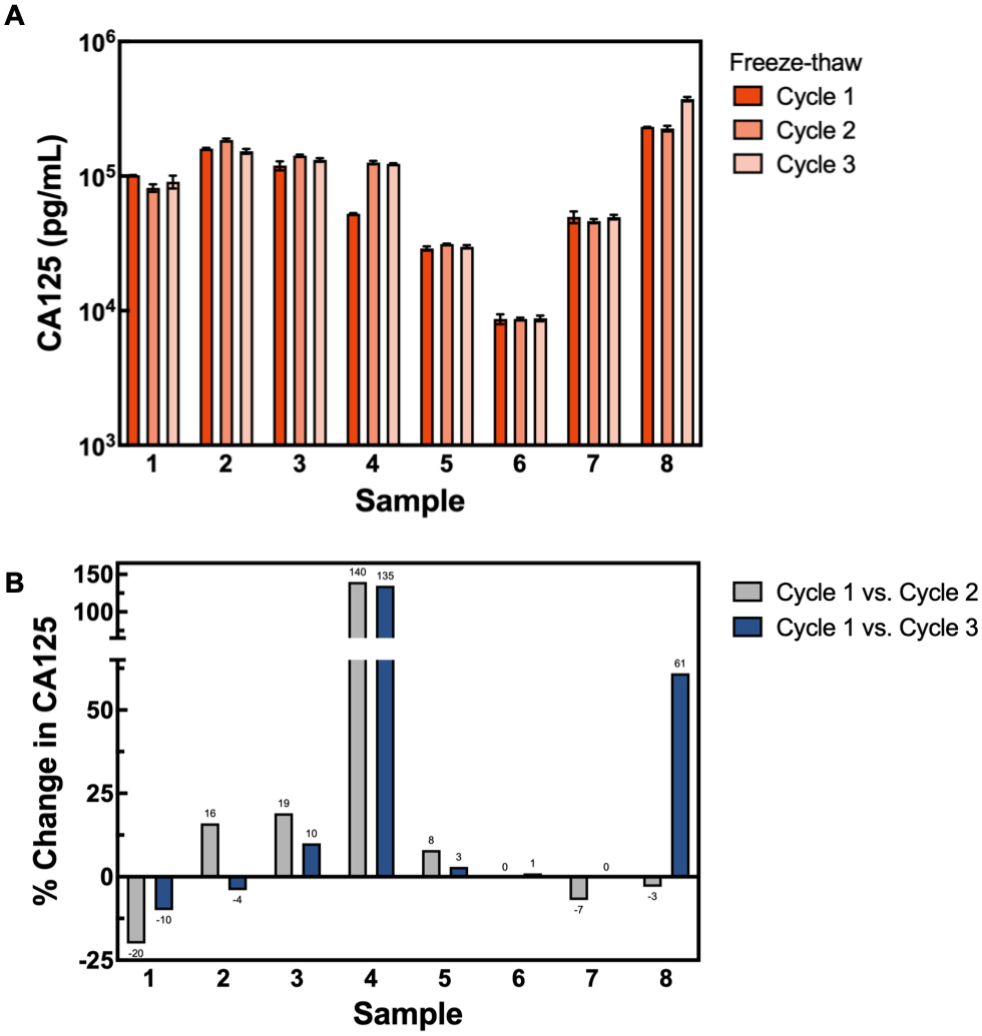
(A) CA125 concentration in a single aliquot of eight plasma samples over three FT cycles (*N* = 8). Data are the average of two replicates, with error bars representing the SD. (B) Percent change in CA125 concentration for each sample compared to the concentration of CA125 at Cycle 1.

### Storage Temperature Impacts Fractionation Success

We investigated how storage conditions affect sample stability by dividing four ME samples into aliquots and storing them at ambient temperature (∼20°C), 4°C, and −20°C for up to 120 hours to span a realistic range of shipping and storage durations (6–120 hours) and temperatures based on priority parcel services in Canada. We also used plasma colour as a qualitative proxy for hemolysis, which we anticipated could interfere with downstream CA125 measurements (Figure 5A). Fractionation success varied markedly by storage condition. ME samples stored at ∼20°C showed a gradual decline in fractionation success over 120 hours, from 100% at 6 hours (4/4 samples studied), to 75% at 24 hours, and 50% by 120 hours (Figure 5B), consistent with progressive sample degradation. Samples stored at 4°C maintained 100% fractionation success over the 120-hour period; however, they also showed visibly increased hemolysis over time. In contrast, ME stored at −20°C failed to fractionate at any time point during the 120-hour period.

**Figure 5.**
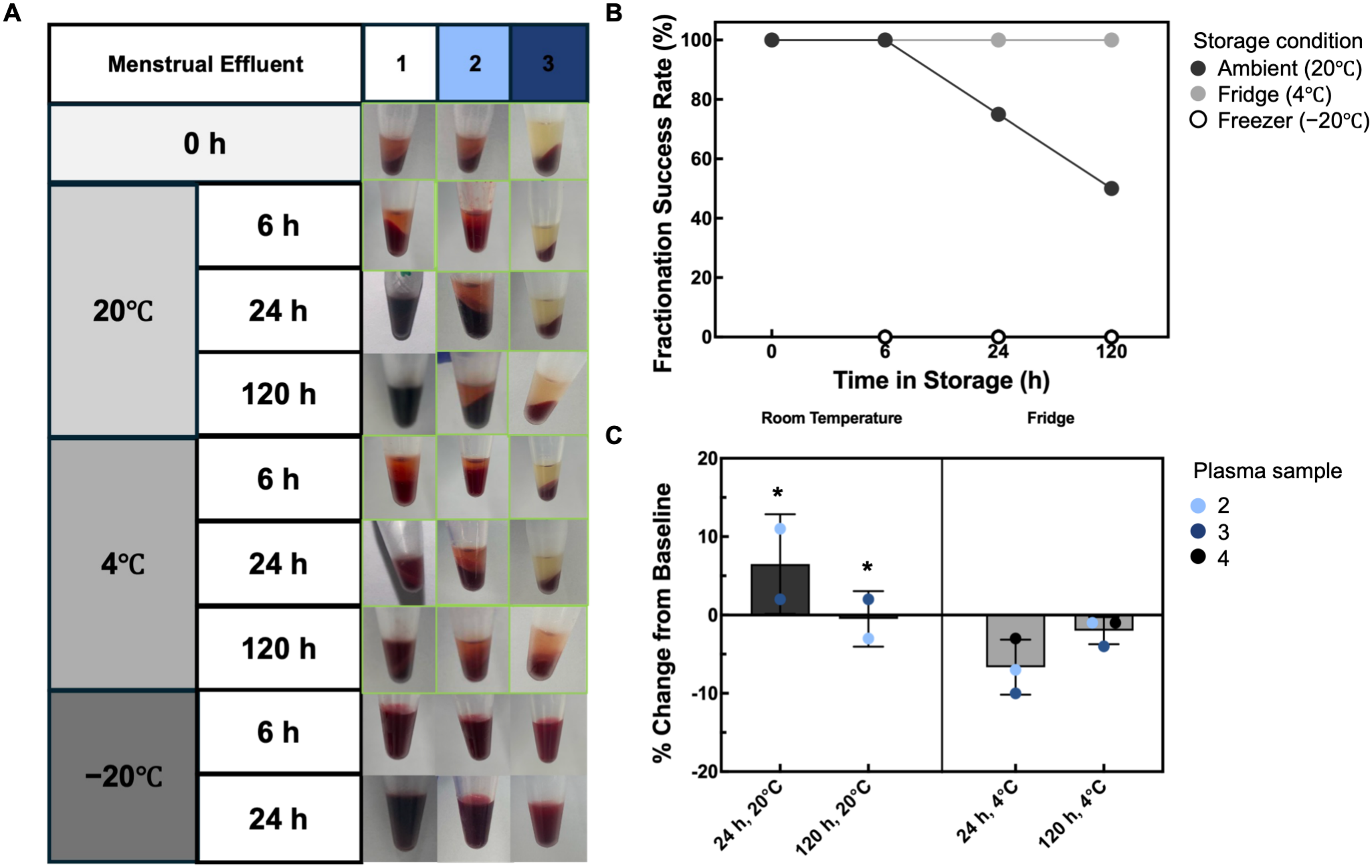
(A) Image of ME samples under different storage conditions after centrifugation (*N* = 3); fractionated samples are boxed in green. (B) Fractionation success by storage condition over time (*N* = 4). (C) Percent CA125 change in fractionated samples under different storage conditions. Data are presented as the mean ± SD. The point marked with (*) indicates sample 4, which fell below acceptance criteria at 24 hours and failed fractionation at 120 hours.

We also measured the CA125 concentration in the plasma that fractionated successfully (Supplemental Table 10). At 24 hours, the change in CA125 was comparable for samples stored at ∼20°C and 4°C (+6.7% and -6.5%, respectively). After 120 hours, only two samples stored at ∼20°C fractionated, with a mean change of -0.5%. The three samples stored at 4°C for 120 hours successfully fractionated, with a mean CA125 change of -2% (Figure 5C). Although the mean change in CA125 levels is comparable between the two conditions, the successful fractionation of all three refrigerated samples indicates that low-temperature storage provides greater storage stability.

### CA125 levels Vary Across the Menstrual Cycle

We quantified CA125 levels in ME samples collected from 19 participants (*N* = 14 ME, *N* = 16 plasma; 9 were matched samples). CA125 was detected in all samples across both matrices (Figure 6A). The average CA125 concentration in ME and plasma was 195,000 ± 238,000 pg/mL (11,000 ± 13,000 U/mL and 158,000 ± 122,000 pg/mL (8,700 ± 6,800 U/mL; see Supplemental Information S4 for the conversion calculation), with plasma showing consistently higher CA125 concentrations than ME in the nine matched samples (Supplemental Figure 4, Supplemental Table 7).

**Figure 6.**
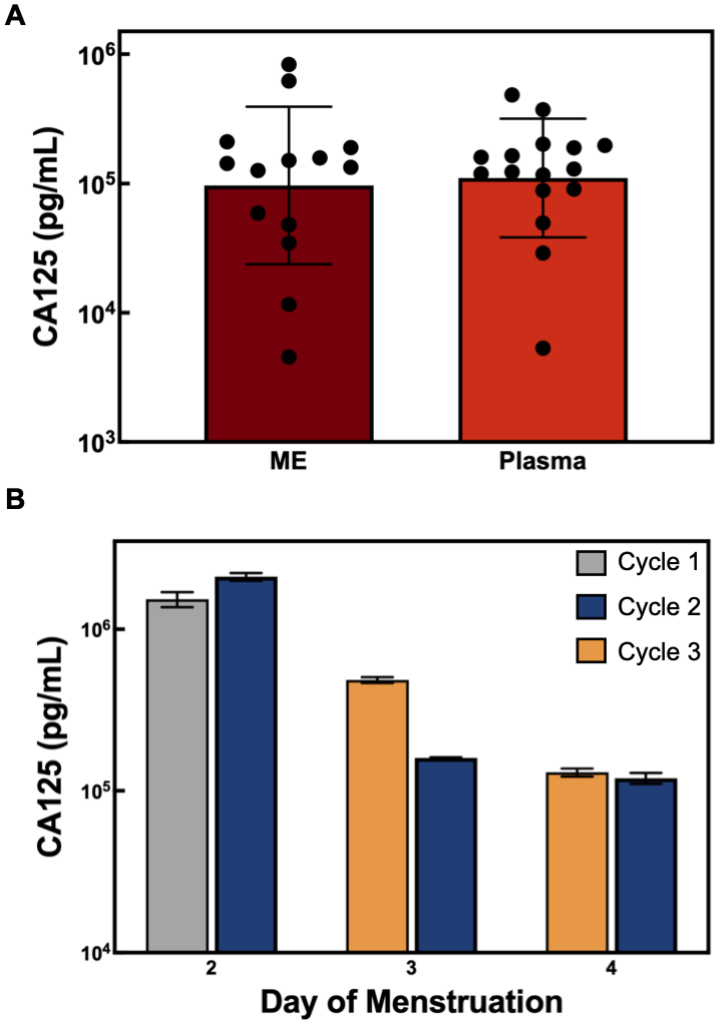
(A) Quantification of CA125 in individual participant samples (*N* = 14 ME, *N* = 16 plasma samples). Each circular symbol represents one sample. (B) Mean CA125 concentration in samples collected on day 2 of the first cycle (grey), days 2-4 of the second cycle (blue), and days 3-4 of the third cycle (yellow). The data are reported as the mean with error bars representing the SD.

To assess the variability in CA125 levels across a menstrual cycle and between cycles, we analyzed ME from one participant who collected samples for the duration of their menstrual cycle and over three cycles (Supplemental Table 11). Consistent with previous serum literature, CA125 concentration in ME collected on three consecutive menstrual days (Figure 6B: blue bars) fluctuates, with the highest CA125 concentration on day 2, followed by a gradual decline on days 3 and 4 of the menstrual cycle (34,35). This trend of declining CA125 levels is also consistent across samples collected across cycles (Figure 6B: grey and yellow bars).

## DISCUSSION

ME has been collected using various methods, including menstrual cups (22,23), pads (24,25), tampons (26), and uterine aspiration (29). We chose menstrual cups for sample collection because they are most commonly used in the literature, allow ME collection with minimal contamination from absorbent materials, and eliminate the need for an additional extraction step before analysis. We used a user-centred approach and conducted user-engagement surveys to design self-sampling kits (Supplemental Information S2). We selected Simoa as our quantification method for its ultrasensitivity, which exceeds conventional ELISA sensitivity by over 1,000-fold, detecting sub-femtomolar concentrations in biofluids such as blood (29), saliva (30), and cerebrospinal fluid (31). Simoa also enables simultaneous detection of multiple biomarkers from a single sample, making it compatible with ME, in which sample collection is limited to menstruation.

Although the median wear time was 3 hours 24 minutes, 35% of samples (*N* = 9) were collected after the cup was worn longer than requested. Some variability likely reflects participants’ lifestyle constraints. However, such deviations can be analytically significant, as extended cup wear may compromise sample integrity (e.g., through biomarker degradation or the release of intracellular components). Notably, as we observed no correlation between the volume of ME collected and the length of cup wear, fixed ME collection windows do not produce consistent ME volumes. As only 10 µL of each sample is needed for Simoa analysis, all samples, irrespective of wear duration, yielded sufficient ME for processing and subsequent analysis. The 3-4-hour collection window should be maintained as a guideline for participants. Future studies should also explicitly report the variation in sample collection times.

Samples that failed to fractionate had longer intervals between sample collection and drop-off (*p* < 0.05), suggesting that processing delays contribute to failed fractionation. We hypothesize that processing delays enable degradation of sample components (e.g., red blood cell lysis or cellular breakdown), which compromises the ability to physically separate plasma from cellular material (33,34). Further investigation is needed to understand the causes of differences in ME’s ability to fractionate. To reduce the risk of fractionation failure, ME should be processed within eight hours of sample collection.

### Plasma Provides Low Aliquot-to-aliquot Variability

Laboratories and biobanks routinely divide biospecimens into aliquots to facilitate storage, enable sample analysis across experiments and laboratories, and minimize FT cycles. These applications assume that each aliquot is representative of the original biospecimen in composition and protein concentration. However, aliquots are not always homogeneous, particularly for complex biological matrices such as tissue biopsies, and faecal samples (35). High aliquot variability can undermine the reliability of protein measurements and compromise the interpretation and comparability of results. No study has explicitly examined inter-aliquot reproducibility in ME. The aliquot-to-aliquot variability observed in ME samples exceeds the intra-assay and inter-assay variability; therefore, differences between ME aliquots are not due to measurement imprecision. ME contain components such as blood clots, endometrial tissue, and cellular material that may not be uniformly distributed throughout the sample. Consequently, a single ME aliquot may not capture the heterogeneity of the original sample. Plasma, however, exhibits low variability across aliquots; therefore, a single aliquot is sufficient to represent a sample’s overall CA125 composition. While anticipated, differences in aliquot-to-aliquot variability between ME and plasma have not been previously quantified. Future testing should investigate methods to reduce the variability observed for ME, such as increased mixing.

### FT cycling Introduces Variability in Plasma CA125 concentrations

Repeated FT cycling has been shown to cause plasma protein aggregation and degradation, altering both biomarker abundance and detectability (36). Plasma samples subjected to three consecutive FT cycles showed inconsistent changes in CA125 concentration. The observed changes likely reflect stress-induced protein instability and the release of intracellular content that alters plasma composition. Such variability can reduce reproducibility and limit clinical translation of protein biomarkers. Therefore, samples should be aliquoted during initial processing, and each aliquot should be limited to a single FT cycle before analysis.

### Storage Temperature Impacts Fractionation Success

For clinical implementation, self-collected ME samples are typically transported from a participant’s home to the laboratory before processing, introducing a period of uncontrolled storage. Fractionation success varied markedly by storage condition. Importantly, the three samples stored at 4°C for 120 hours before undergoing CA125 quantification successfully fractionated, with a mean CA125 change of -2%. We therefore recommend refrigeration (4°C) as the preferred storage condition following ME self-collection and during transport to the laboratory to improve sample stability. In future studies, samples should be transported with ice packs to more closely approximate 4°C conditions.

### CA125 levels Vary Across the Menstrual Cycle

The average plasma CA125 concentration of 9,500 ± 7,600 U/mL was lower than the 47,000 ± 35,000 U/mL reported by Takahashi et al., who investigated the use of CA125 levels in ME for distinguishing patients with and without endometriosis (14). The discrepancy in CA125 levels likely reflects preanalytical differences. Takahashi et al. analyzed vaginal aspirated ME collected on day 3 from 30 controls evaluated or treated for infertility or pelvic pain. These conditions may elevate CA125 baseline levels, despite the patients’ suitability as endometriosis controls. Our findings show that CA125 can be quantified in ME and plasma collected from 19 participants using a minimally invasive menstrual cup.

Changes in CA125 levels over the menstrual cycle indicate that the day of sample collection matters, as CA125 levels can vary by up to 92% (roughly one order of magnitude) between days 2 and 3. If the day of sample collection is uncontrolled, CA125 fluctuations may be misinterpreted as biological differences between individuals. Given that the data were derived from samples provided by one participant, these preliminary findings warrant further investigation in a larger cohort. Nevertheless, self-collected ME successfully revealed variations in CA125 concentration across a participant’s menstrual cycle, demonstrating the feasibility of longitudinal monitoring of protein biomarkers over a menstrual cycle.

### Recommended Workflow for Protein Detection in ME

Based on this report, we recommend the following guidelines for collecting and handling ME for protein biomarker analysis, with an expanded protocol available in Supplementary Information S5. Participants should collect ME on a defined menstruation day within a 3–4-hour window. The collected ME should be transferred to a K_2_EDTA blood collection tube and returned to the laboratory within eight hours of sample collection. Preliminary data suggest that at-home storage and transportation of ME at 4 °C could preserve sample stability for up to 120 hours. Process samples immediately after drop-off; if immediate processing is not possible, refrigerate at 4 °C.

We had no consensus on whether ME or plasma is the superior biofluid for protein biomarker discovery; therefore, choose ME or plasma based on the study objectives. For plasma proteomic studies, the ME sample can be centrifuged at 2,000 × g at 4°C for 30 minutes. If the sample fractionates, carefully collect the plasma-like layer in aliquots. Both ME and plasma should be stored at −80°C promptly after processing for at least 24 hours prior to analysis. Additionally, each aliquot should undergo no more than one FT cycle. Documenting ME processing ensures standardized, reproducible procedures and reduces preanalytical variability. Record key parameters (e.g., collection day, cup wear duration, pre-centrifugation delay duration), as they can influence sample stability and subsequent biomarker detection and interpretation.

## CONCLUSION

In this work, we successfully quantified CA125, a model protein biomarker, in menstrual cup-collected ME and in plasma using Simoa technology. We show that preanalytical factors, including FT cycling and storage conditions, affect CA125 concentration. We preliminarily demonstrate that the developed workflow can be used for longitudinal monitoring of CA125. Finally, we provide guidelines to support the development of a standardized workflow for protein analysis in ME. This study has several limitations. First, the data are limited by a small sample size (*N* = 26 total samples). Notably, 19% of samples failed to fractionate. While processing delays may contribute to failed fractionation (*p* < 0.05), two samples stored at ambient temperature remained stable for 120 hours. Future studies should examine the inter-person compositional variations that may contribute to differences in sample stability. Extensive work is still required to optimize other preanalytical parameters, such as collection tube type, centrifugation conditions, and long-term storage conditions. Despite using low-temperature, low-speed centrifugation, we observed varying degrees of hemolysis across all samples. Optimizing centrifugation parameters, including duration, speed, and temperature, could enable faster processing and reduce hemolysis. Furthermore, all results presented are based on CA125. The results in this report should be validated using additional protein biomarkers to confirm the method’s robustness for protein detection in ME. Future work will involve assessing the clinical significance of ME CA125 in gynecological diseases, including endometriosis and ovarian cancer, by comparing CA125 concentrations in cohorts with and without these diseases. The developed workflow supports the use of self-collected ME as an accessible, minimally invasive biofluid suitable for protein biomarker quantification towards gynecological disease diagnosis.

## Data Availability

All data produced in the present study are available upon reasonable request to the authors.

## ASSOCIATED CONTENT

Supporting Information.

### Author Contributions

Shekemi Denuga: Conceptualization, formal analysis, investigation, methodology, visualization, writing (original draft).

Barbara Lukaszczyk: Conceptualization, formal analysis, investigation, methodology, visualization, writing (review & editing).

Theresa Okwuegbuna: Conceptualization, investigation, writing (review & editing).

Kezia E. Suryoraharjo: Conceptualization, investigation, writing (review & editing).

Alana F. Ogata: Conceptualization, supervision, funding acquisition, project administration, writing (review & editing).

## Acknowledgments

The authors thank all participants who completed the online surveys or self-collected menstrual effluent. Without your participation, we would not have been able to complete this study. We also wish to acknowledge the land on which the University of Toronto operates. For thousands of years, it has been the traditional land of the Huron-Wendat, the Seneca, and the Mississaugas of the Credit. Today, this meeting place is still the home to many Indigenous people from across Turtle Island, and we are grateful to have the opportunity to work on this land.

