## Supplemental Information for "Rethink the Flow: Establishing a Preanalytical Workflow for Protein Detection in Menstrual Effluent"

Shekemi Denuga,[1,2] Barbara Lukaszczyk,[1,2] Theresa Okwuegbuna,[1,2] Kezia E. Suryoraharjo[1,2] Alana F. Ogata.[1,2]*

1. Department of Chemistry, University of Toronto, Toronto, ON, Canada.

2. Department of Chemical and Physical Sciences, 2University of Toronto Mississauga, Mississauga, ON, Canada.

*alana.ogata @utoronto.ca

### S1. Supplemental Methods

#### S1.1 User Engagement

We conducted a pre-study survey of 50 individuals aged 18 years and older to understand public perceptions of menstrual cups and to collaboratively develop an ME self-collection protocol and self-collection kit suitable for end users (1). Survey participants were recruited via social media, physical flyers, and word of mouth under a protocol approved by the University of Toronto Research Ethics Board (Human Protocol #00048901). All participants provided written informed consent.

#### S1.2 Simoa Assay

All CA125 measurements were performed using a single-molecule array (Simoa) assay on the HD-X Analyzer (Quanterix). The CA125 Simoa assay used for CA125 characterization in ME was developed in-house as previously reported (2), using the CA125 assay kit purchased from R&D Systems (cat# DY5609-05).

##### *S1.2.1 Conjugation of CA125 Capture Antibodies to Paramagnetic Beads*

The CA125 capture antibody was conjugated to carboxylated paramagnetic beads according to the Simoa Bead-based Homebrew Assay Development Guide from Quanterix. First, the antibodies were buffer-exchanged into bead conjugation buffer using an Amicon centrifugal filter. The filter was centrifuged at 14,000 × g for 5 minutes; the flow-through was discarded, and the process was repeated twice. To recover the antibody, the filter was inverted into a new Amicon microcentrifuge tube and centrifuged at 1,000 × g for 2 minutes. 50 µL of Bead Conjugation Buffer was added to the filter before centrifuging again at 1,000 × g for 2 minutes. The concentration of the buffer-exchanged antibody was measured using a spectrophotometer at OD 280 (Thermo Fisher). The antibody was then diluted to 0.2 mg/mL using Bead Conjugation Buffer and stored on ice until ready to use. The carboxylated paramagnetic beads were washed three times with Bead Wash Buffer and adjusted to a concentration of $1.4 \times 10^9$ beads/mL. The beads were subsequently washed twice with Bead Conjugation Buffer and resuspended in ice-cold Bead Conjugation Buffer. The beads were activated with 10 mg/mL 1-ethyl-3-(3-dimethylaminopropyl) carbodiimide hydrochloride (EDC) (Thermofisher) to a final concentration of 0.3 mg/mL. The beads were activated at 4°C for 30 minutes on a rotator. The antibody-conjugated beads were then washed twice with Bead Wash Buffer and blocked by rotating in Bead Blocking Buffer for 45 minutes at 4°C. Finally, the beads were washed twice with Bead Wash Buffer and resuspended in Bead Diluent.

*S1.2.2 CA125 Measurements*

The prepared capture antibody-conjugated beads were diluted to 2 x $10^7$ beads/mL in Bead Diluent. The biotinylated detector antibody was reconstituted according to the manufacturer's instructions and diluted to 0.1 µg/mL in Homebrew Detector/Sample Diluent. The streptavidin β-galactosidase (SβG) Concentrate was diluted to 150 pM in SβG Diluent. The CA125 assay was performed using a three-step protocol. 25 µL of the bead solution and 100 µL of sample were incubated for 15 minutes, followed by a wash with System Wash Buffer 1. The beads were resuspended in 100 µL of biotinylated detector antibody and incubated for 5 minutes 15 seconds, followed by a wash with System Wash Buffer 1. The beads were then resuspended in 100 µL of SβG and incubated for 5 minutes 15 seconds, followed by six washes with System Wash Buffer 1 and a final wash with System Wash Buffer 2. The beads were resuspended in 25 µL of Resorufin-β-D-galactopyranoside (RGP) Reagent before being loaded into the Simoa disc. The microwell array was sealed with Simoa Sealing Oil and analyzed by imaging. Average Enzyme per Bead (AEB) values were fit against biomarker concentrations using a 4-parameter logistic curve weighted by $1/y^2$ to generate calibration curves. Each independent Simoa run included nine calibration levels to generate a calibration curve. The calibrators were prepared by serial dilution of recombinant protein standards in Homebrew Detector/Sample diluent, ranging from 0.41 pg/mL to 900 pg/mL. Three quality control samples at high (540 pg/mL), medium (54 pg/mL), and low (5.4 pg/mL) concentrations, prepared in Homebrew Detector/Sample Diluent, were also included in each run, with acceptable error margins of ±20%. Menstrual effluent (ME) samples were diluted in Homebrew Detector/Sample Diluent. All calibrators and samples were measured in duplicate, with CA125 levels for each sample determined using the calibration curve prepared on the same day as the analysis.

*S1.2.3 CA125 Assay Validation*

Prior to CA125 measurements, ME and plasma samples were thawed on ice, vortex-mixed for 30 seconds and centrifuged briefly using a mini centrifuge. Pooled samples were prepared by combining 10 µL from at least three participants' samples, then vortexing and centrifuging. For dilution linearity, the pooled sample was serially diluted with Homebrew Detector/Sample Diluent and analyzed by Simoa. Spike-and-recovery experiments were also conducted using pooled samples and eight individual samples. Each sample was analyzed unspiked and after spiking with low (2 pg/mL), medium (10 pg/mL), and high (100 pg/mL) concentrations of recombinant CA125. Percent recovery at each spike level was calculated using equation 1, with an acceptance criterion of 70-130%.

$$\%\,recovery = \left(\frac{Spiked\ concentration - unspiked\ concentration}{expected\ spike\ concentration}\right) x\ 100 \quad \textbf{Eq. 1}$$

#### *S1.2.4 CA125 Quantification in ME and Plasma*

Prior to the Simoa measurement, each sample was thawed on ice, vortexed for 30 seconds, and briefly centrifuged to ensure homogeneity. ME samples were diluted 60,000-fold, and plasma samples were diluted 8,000-fold in Homebrew Detector/Sample Diluent. The accuracy of CA125 concentration in each sample was validated by performing spike-and-recovery, in which each sample was analyzed unspiked and after spiking with 2 pg/mL, 10 pg/mL, and 100 pg/mL of recombinant CA125. Percent recovery at each spike level was calculated using equation 1. The average percent recovery across the three spike levels was then calculated for each sample with an acceptance criterion of 70-130%. Samples that fell below the acceptable range of 70–130% were excluded from this study. To quantify the total CA125 concentration in the sample, we multiplied the concentration obtained by Simoa by the dilution factor.

### S2. User Engagement

#### S2.1 Pre-study User Engagement

We used an iterative, user-centred approach to design self-sampling kits. First, we assessed public attitudes toward ME self-collection using menstrual cups (Supplemental Table 1). Of 50 respondents, 22% had prior experience using menstrual cups. However, 32% reported they would be willing to use a menstrual cup for the study. This growing interest in menstrual cups is consistent with previous studies that have also reported their rise in popularity, largely because they are environmentally friendly (3,4). 42% of people surveyed said they would collect samples only at home, while 56% would feel comfortable collecting a sample either at home or in the bathroom near the laboratory. Another 4% would not feel comfortable self-sampling. Therefore, to reduce barriers to menstrual cup use, we developed a self-sampling protocol compatible with at-home collection. 62% of survey participants felt comfortable using commercially available menstrual products provided by the lab, while 40% preferred to use their own. We concluded that participants prefer the laboratory to provide one menstrual cup. The final self-sampling kits included one menstrual cup, a pair of disposable gloves, one blood collection tube, a biohazard bag, a menstrual cup protocol, a sampling information card, and a QR code linking to a post-collection survey. A picture of the collection kit, sample collection protocol and sample information card can be found in Supplemental Figure 1. Survey respondents also received the menstrual cup protocol to assess its comprehensibility and usability. 98% reported confidence in their ability to follow the instructions to self-collect a sample, and 92% agreed or strongly agreed that they could follow the instructions without a physical demonstration, such as a video. Based on feedback from the general public survey and the post-sample collection survey of participants who self-sampled, we made 3 rounds of changes to the protocol. Changes included bolding key information in the protocol, adding alternative menstrual cup folding techniques for successful insertion, and expanding the collection day from the second day of the menstrual cycle to the day of heaviest flow. Through this iterative design process, we developed an accessible self-sampling kit with a standardized protocol for collecting ME at home; 100% of participants who self-sampled agreed or strongly agreed that using the kit would be more convenient than visiting a clinic for diagnostic testing (Supplemental Table 3).

**Supplemental Table 1:** Pre-study user engagement survey

| Question | *N* participants | % |
|---|---|---|
| Have you ever used a menstrual cup, pad, or tampon before? (Select all that apply) | | |
| Pad | 50 | 100% |
| Tampon | 38 | 76% |
| Cup | 11 | 22% |
| None | 0 | 0% |
| Which product would you most likely use if you were to participate in the study? (Select all that apply) | | |
| Pad | 30 | 60% |
| Tampon | 22 | 44% |
| Cup | 16 | 32% |
| None | 0 | 0% |
| Would you be comfortable with using a commercially available menstrual product provided by the lab in the collection kit, or would you prefer to use your own product for sample collection (you would need to let the lab know what company the product is from)? | | |
| Lab provide | 30 | 60% |
| My own | 20 | 40% |
| Would you feel comfortable collecting your sample in either a home setting or a campus bathroom? | | |
| Only at home | 21 | 42% |
| Either at home or on campus | 27 | 54% |
| None of the above, I am not comfortable with self-sampling | 2 | 4% |
| I am confident that I could follow these instructions to self-collect a sample. | | |
| Strongly agree | 31 | 62% |
| Agree | 17 | 34% |
| Neutral | 1 | 2% |
| Disagree | 1 | 2% |
| It was easy to follow the step-by-step instructions without any physical demonstration (e.g. a video). | | |
| Strongly agree | 29 | 58% |
| Agree | 16 | 32% |
| Neutral | 3 | 6% |
| Disagree | 2 | 4% |
| The instructions clearly explain how to avoid contamination or errors. | | |

| | | |
|---|---|---|
| Strongly agree | 28 | 56% |
| Agree | 14 | 28% |
| Neutral | 5 | 10% |
| Disagree | 3 | 6% |
| The self-sampling kit provided clear instructions that only one tampon/pad/cup is needed, and I did not feel the need for an extra tampon/pad/cup. | | |
| Strongly agree | 28 | 56% |
| Agree | 11 | 22% |
| Neutral | 8 | 16% |
| Disagree | 3 | 6% |
| The layout of the instructions was easy to read. | | |
| Strongly agree | 33 | 66% |
| Agree | 13 | 26% |
| Neutral | 4 | 8% |
| Disagree | 0 | 0% |
| A 24-hour timeframe would be sufficient for me to drop off my sample at the Ogata Lab on the UTM campus. | | |
| Strongly agree | 20 | 40% |
| Agree | 10 | 20% |
| Neutral | 9 | 18% |
| Disagree | 7 | 14% |

### S2.2 Self-sampling Participant Engagement and ME Collection

Each participant received a self-collection kit containing a menstrual cup (sizes small/large available), a $K_2EDTA$ blood collection tube, a biosafety hazard bag, a pair of gloves, a menstrual cup protocol, a sampling info card for completion, and a QR code for a post-collection survey (Supplemental Figure 1).

A B

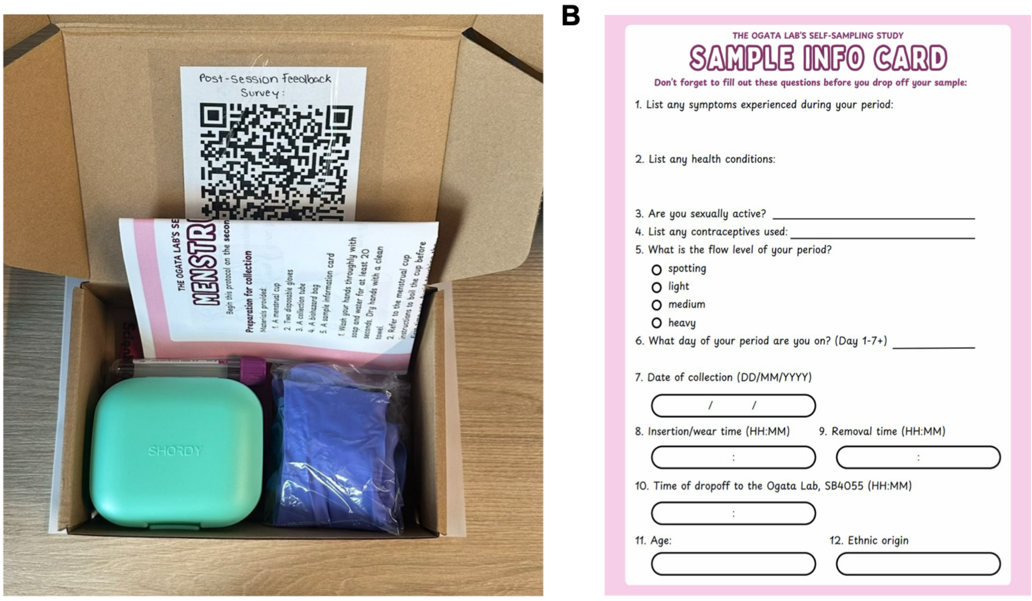

**Supplemental Figure 1:** (A) Photo of self-collection kits provided to participants. (B) The sample information card given to participants to record details about their health, demographics, and the sample collection process.

**Supplemental Table 2:** Demographic characteristics of study participants.

| **Characteristic** | **Result** |
|---|---|
| Participants, *N* | 19 |
| Samples, *N* | 26 |
| Age, years, median (IQR; range), *N* = 11 | 25 (21–34; 19–40) |
| Ethnic origin (self-described), *N* participants | |
| Pakistani | 1 |
| Indian | 1 |
| Jewish | 1 |
| Asian | 1 |
| Brazil | 1 |
| Japanese | 1 |
| Middle Eastern | 1 |
| Caucasian | 2 |
| Mixed Race | 1 |
| Not recorded | 9 |

| Characteristic | Result |
|---|---|
| Hormonal Contraception: | 2 |
| Health condition (self-reported), *N* participants | 4 (fibroids; PCOS; type 1 diabetes/lupus; pituitary mass) |
| Self-reported flow: light / medium / heavy, *N* samples | 7 / 13 / 6 |
| Day of period: 1 / 2 / 3 / 4 / 5, *N* samples | 2 / 13 / 5 / 5 / 1 |
| Fractionation Rate: Successful / Failed, *N* samples | 21 / 5 |
| Mild hemolysis | 4 |
| Moderate hemolysis | 10 |
| Severe hemolysis | 7 |
| Not applicable | 5 |

**Supplemental Table 3:** Results from post-collection survey

| Question | N Participants | % |
|---|---|---|
| Instructions provided were easy to follow | | |
| Strongly Agree | 8 | 50% |
| Agree | 6 | 38% |
| Neutral | 1 | 6% |
| Disagree | 1 | 6% |
| The self-sampling kit provided clear instructions that only one pad/tampon /cup was needed, and I did not feel the need for an extra pad/tampon/cup. | | |
| Strongly Agree | 8 | 89% |
| Agree | 1 | 11% |
| Neutral | 0 | 0% |
| Disagree | 0 | 0% |
| I am confident that I was able to avoid contamination or errors. | | |
| Strongly Agree | 4 | 25% |
| Agree | 8 | 50% |
| Neutral | 2 | 13% |
| Disagree | 0 | 0% |
| The 24-hour timeframe was sufficient for me to drop off my sample at the Ogata Lab on the UTM campus. | | |
| Strongly Agree | 9 | 56% |
| Agree | 6 | 38% |
| Neutral | 0 | 0% |
| Disagree | 1 | 6% |
| I am comfortable with collecting my sample on the go in a non-home setting (e.g., a public restroom). | | |
| Strongly Agree | 2 | 13% |

| | | |
|---|---|---|
| Agree | 1 | 6% |
| Neutral | 2 | 13% |
| Disagree | 8 | 50% |
| I would prefer to use my regular menstrual product (e.g., preferred brand, size, type) rather than the commercially available product provided by the lab in the self-sampling kit. | | |
| Strongly Agree | 1 | 11% |
| Agree | 2 | 22% |
| Neutral | 4 | 44% |
| Disagree | 2 | 22% |
| I believe that collecting menstrual blood using an at-home self-sampling kit would be more convenient than going to a clinic or undergoing other diagnostic testing (e.g., urine sample or venous blood draw) . | | |
| Strongly Agree | 6 | 38% |
| Agree | 10 | 63% |
| Neutral | 0 | 0% |
| Disagree | 0 | 0% |
| I would trust the accuracy of the test result from a self-collected menstrual blood sample. | | |
| Strongly Agree | 2 | 13% |
| Agree | 10 | 63% |
| Neutral | 4 | 25% |
| Disagree | 0 | 0% |
| I would be willing to collect my own sample using the self-sampling kit during each menstrual cycle or as needed. | | |
| Strongly Agree | 5 | 31% |
| Agree | 10 | 63% |
| Neutral | 1 | 6% |
| Disagree | 0 | 0% |
| I would be open to using a different type of menstrual product (pad/tampon/cup) for self-sampling in the future. | | |
| Strongly Agree | 6 | 38% |
| Agree | 7 | 44% |
| Neutral | 1 | 6% |
| Disagree | 0 | 0% |

### S3. Simoa CA125 Measurements

#### S3.1 Validation of CA125 Assay

The CA125 assay achieved a limit of detection of 0.1 ± 0.1 pg/mL ($N$ = 19) with an inter-assay variability of 6% and intra-assay variability of 4% (Supplemental Figure 2, Supplemental Table 4-6).

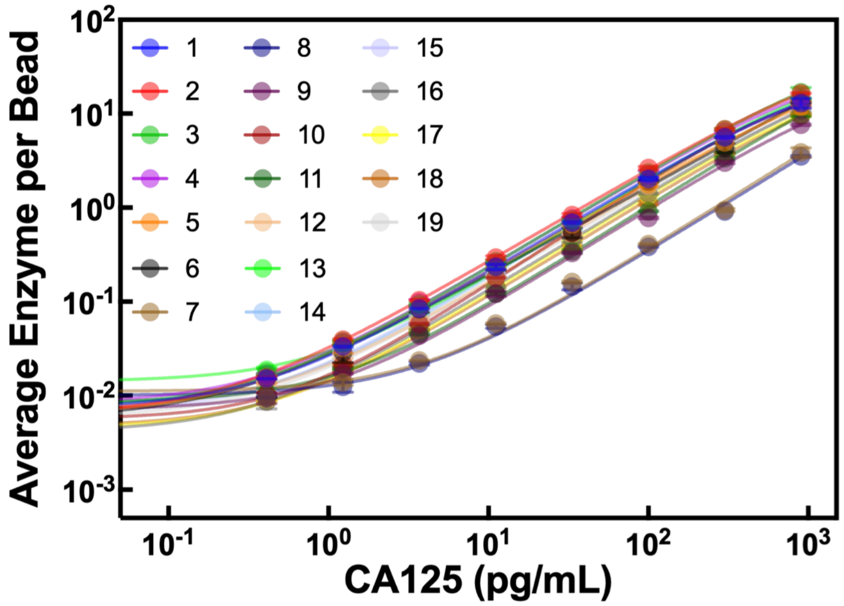

**Supplemental Figure 2:** Simoa calibration curve plot from 19 independent runs.

**Supplemental Table 4:** Analytical parameters of CA125 calibration curves (*N* = 19).

| LOD (pg/mL) | Blank signal (AEB) | SD of first non-blank calibrator (AEB) | 4PL fit coefficient A | 4PL fit coefficient B | 4PL fit coefficient C | 4PL fit coefficient D |
|---|---|---|---|---|---|---|
| 0.00550 | 0.00662 | 0.00004 | 0.00663 | 0.99991 | 1985.73461 | 41.89563 |
| 0.06000 | 0.00681 | 0.00044 | 0.00669 | 1.01542 | 1689.72531 | 47.30614 |
| 0.15747 | 0.00548 | 0.00144 | 0.00559 | 0.97627 | 6776.39218 | 140.70614 |
| 0.04010 | 0.00690 | 0.00030 | 0.00690 | 0.99066 | 2537.80678 | 57.48142 |
| 0.20790 | 0.00620 | 0.00157 | 0.00615 | 0.97086 | 2518.53642 | 43.79377 |
| 0.01012 | 0.00583 | 0.00007 | 0.00579 | 0.95155 | 22675.62397 | 292.68001 |
| 0.00597 | 0.00695 | 0.00001 | 0.00710 | 0.94145 | 784660498.15903 | 1233805.62114 |
| 0.05414 | 0.00738 | 0.00010 | 0.00741 | 0.96097 | 1868184.06600 | 4711.13494 |
| 0.30077 | 0.00415 | 0.00120 | 0.00452 | 1.07669 | 1496.70890 | 35.20608 |
| 0.14632 | 0.00441 | 0.00060 | 0.00452 | 1.07669 | 1496.70890 | 35.20608 |
| 0.33660 | 0.00585 | 0.00125 | 0.00600 | 0.99404 | 44880705265789.60000 | 392875823181.25000 |
| 0.13930 | 0.00649 | 0.00075 | 0.00851 | 0.98626 | 3269.74483 | 61.21504 |
| 0.23042 | 0.01223 | 0.00138 | 0.01214 | 1.01000 | 3541.86555 | 71.70765 |
| 0.20010 | 0.00563 | 0.00121 | 0.00571 | 1.01666 | 2518.59430 | 52.40764 |
| 0.05899 | 0.00475 | 0.00043 | 0.00487 | 1.00004 | 2528.00282 | 50.47043 |

**Supplemental Table 5:** Forty-five samples of known concentration were tested in separate assays to assess inter-assay variability.

| Sample | Result 1 | Result 2 | Mean (pg/mL) | SD | %CV |
|---|---|---|---|---|---|
| 1 | 150.292 | 142.451 | 146.371 | 5.544 | 4% |
| 2 | 135.636 | 122.328 | 128.982 | 9.410 | 7% |
| 3 | 151.447 | 147.490 | 149.468 | 2.798 | 2% |
| 4 | 141.578 | 146.690 | 144.134 | 3.615 | 3% |
| 5 | 149.022 | 107.293 | 128.157 | 29.507 | 23% |
| 6 | 138.418 | 183.786 | 161.102 | 32.080 | 20% |
| 7 | 129.127 | 127.020 | 128.073 | 1.490 | 1% |
| 8 | 134.264 | 127.592 | 130.928 | 4.718 | 4% |
| 9 | 119.293 | 128.212 | 123.753 | 6.306 | 5% |
| 10 | 111.741 | 106.118 | 108.929 | 3.976 | 4% |
| 11 | 156.277 | 140.659 | 148.468 | 11.043 | 7% |
| 12 | 134.153 | 122.645 | 128.399 | 8.137 | 6% |
| 13 | 135.606 | 141.809 | 138.708 | 4.386 | 3% |
| 14 | 156.277 | 140.659 | 148.468 | 11.043 | 7% |
| 15 | 139.159 | 140.920 | 140.040 | 1.245 | 1% |
| 16 | 15.193 | 14.355 | 14.774 | 0.593 | 4% |
| 17 | 14.123 | 13.624 | 13.873 | 0.353 | 3% |
| 18 | 13.660 | 13.269 | 13.464 | 0.277 | 2% |
| 19 | 13.335 | 12.945 | 13.140 | 0.276 | 2% |
| 20 | 15.126 | 14.108 | 14.617 | 0.720 | 5% |
| 21 | 16.985 | 14.825 | 15.905 | 1.528 | 10% |
| 22 | 13.849 | 13.363 | 13.606 | 0.344 | 3% |
| 23 | 14.028 | 14.439 | 14.234 | 0.291 | 2% |
| 24 | 14.966 | 11.892 | 13.429 | 2.174 | 16% |
| 25 | 14.411 | 15.346 | 14.878 | 0.662 | 4% |
| 26 | 13.846 | 13.006 | 13.426 | 0.594 | 4% |
| 27 | 12.578 | 12.845 | 12.711 | 0.188 | 1% |
| 28 | 12.024 | 12.792 | 12.408 | 0.543 | 4% |
| 29 | 13.846 | 13.006 | 13.426 | 0.594 | 4% |
| 30 | 12.348 | 12.187 | 12.267 | 0.114 | 1% |
| 31 | 1.534 | 1.322 | 1.428 | 0.149 | 10% |
| 32 | 1.312 | 1.465 | 1.389 | 0.108 | 8% |
| 33 | 1.174 | 1.147 | 1.161 | 0.019 | 2% |

| | | | | | |
|---|---|---|---|---|---|
| 34 | 1.341 | 1.343 | 1.342 | 0.001 | 0% |
| 35 | 1.418 | 1.256 | 1.337 | 0.114 | 9% |
| 36 | 1.587 | 1.259 | 1.423 | 0.232 | 16% |
| 37 | 1.542 | 1.558 | 1.550 | 0.012 | 1% |
| 38 | 1.297 | 1.242 | 1.270 | 0.039 | 3% |
| 39 | 1.457 | 1.638 | 1.548 | 0.128 | 8% |
| 40 | 1.457 | 1.638 | 1.548 | 0.128 | 8% |
| 41 | 1.480 | 1.220 | 1.350 | 0.184 | 14% |
| 42 | 1.152 | 1.259 | 1.206 | 0.076 | 6% |
| 43 | 4.245 | 4.447 | 4.346 | 0.143 | 3% |
| 44 | 1.480 | 1.220 | 1.350 | 0.184 | 14% |
| 45 | 1.317 | 1.227 | 1.272 | 0.064 | 5% |
| Average coefficient of variation (CV) | | | | | 6% |

**Supplemental Table 6:** Fourteen samples were tested to assess intra-assay variability.

| Sample | Result 1 | Result 2 | Mean (pg/mL) | SD | %CV |
|---|---|---|---|---|---|
| 1 | 12.63 | 12.73 | 12.68 | 0.07 | 1% |
| 2 | 10.47 | 12.24 | 11.36 | 1.25 | 11% |
| 3 | 21.86 | 22.86 | 22.36 | 0.71 | 3% |
| 4 | 25.28 | 24.06 | 24.67 | 0.86 | 4% |
| 5 | 6.03 | 6.37 | 6.20 | 0.24 | 4% |
| 6 | 45.37 | 47.88 | 46.63 | 1.78 | 4% |
| 7 | 6.62 | 6.50 | 6.56 | 0.09 | 1% |
| 8 | 15.51 | 15.28 | 15.39 | 0.16 | 1% |
| 9 | 0.61 | 0.71 | 0.66 | 0.07 | 11% |
| 10 | 0.64 | 0.69 | 0.67 | 0.04 | 6% |
| 11 | 10.50 | 11.67 | 11.08 | 0.83 | 7% |
| 12 | 11.46 | 11.58 | 11.52 | 0.08 | 1% |
| 13 | 33.93 | 35.60 | 34.77 | 1.18 | 3% |
| 14 | 24.69 | 25.87 | 25.28 | 0.83 | 3% |
| Average CV | | | | | 4% |

Next, we validated the CA125 assay in pooled ME and plasma samples ($N$ = 4 and 3, respectively) using dilution linearity and spike-and-recovery experiments. For dilution linearity, pooled ME or plasma samples were serially diluted, and the Simoa response was measured (Supplemental Figure 3A-

B). The linear dilution range for ME was 10,000x to 220,000x ($R^2 > 0.99$), whereas plasma showed linearity from 3,000x to 30,000x ($R^2 > 0.99$).

We then performed spike-and-recovery experiments using pooled ME and plasma samples spiked with low, mid, and high concentrations of recombinant CA125 protein (2, 10 and 100 pg/mL, respectively). We performed the pooled spike-and-recovery experiment at three dilution factors within the established linear range for pooled ME and plasma samples to assess their compatibility for CA125 quantification (Supplemental Figure 3C-D). For ME and plasma, all three dilution factors yielded percent recoveries within the acceptance range of 70 - 130%. We selected a 60,000x dilution for ME samples because it was closest to 100% recovery while requiring the least dilution. For plasma samples, we selected 8,000x as the optimal dilution factor for the same reason.

Finally, spike-and-recovery experiments were performed on eight individual ME and plasma samples using the optimal dilution factors identified (Supplemental Figure 3E-F). Percent recoveries for all samples fell within the 70–130% range, except for the low spike concentration in plasma sample 1 (133.7%). Variability in percent recovery likely reflects differences in ME matrix composition, with more pronounced effects at the low spike concentration. Although further dilution may improve recoveries, we proceeded with dilution factors of 8000x for plasma and 60,000x for ME, as these yielded detectable CA125 concentrations while minimizing matrix effects.

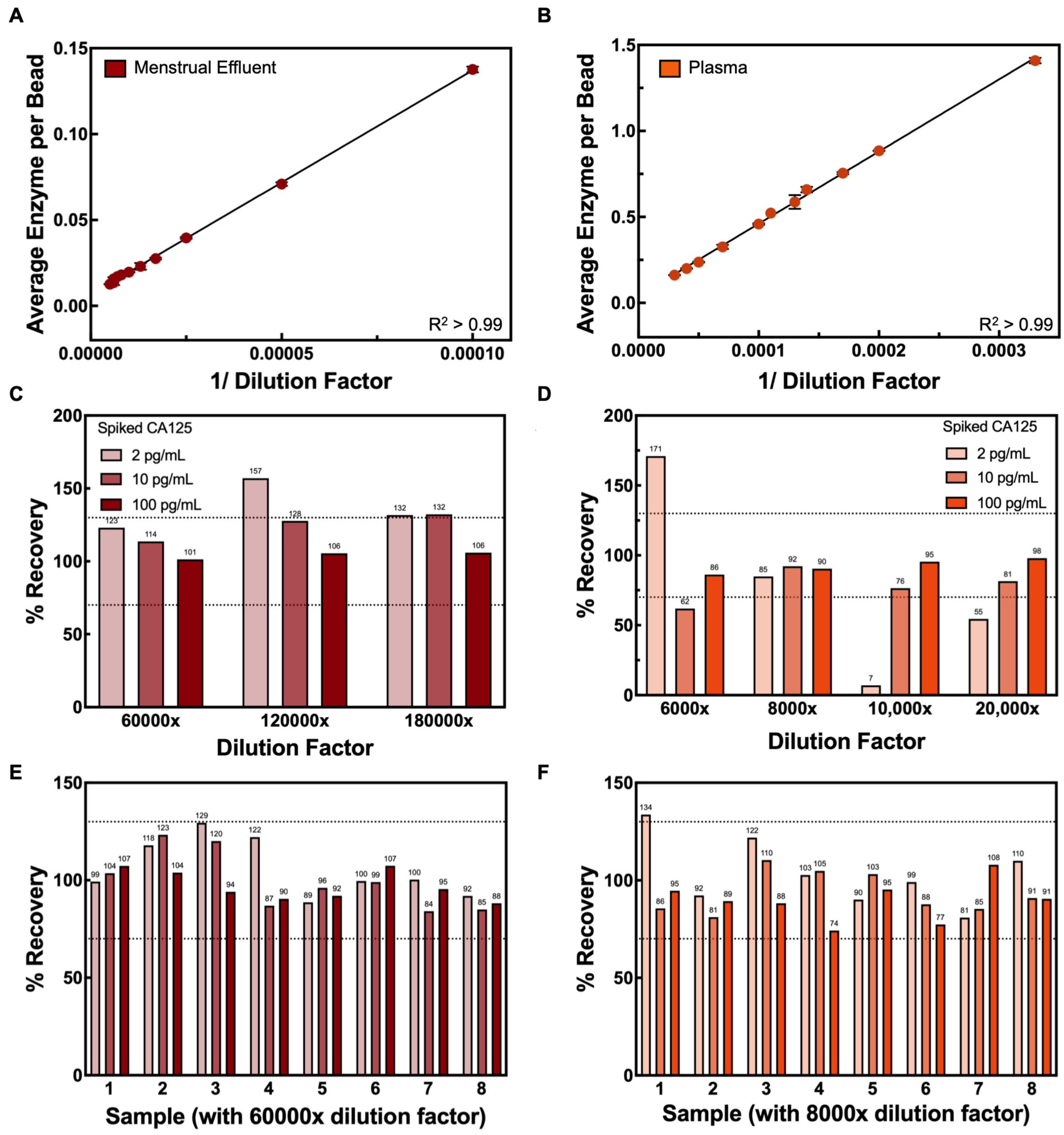

**Supplemental Figure 3:** CA125 assay response over (A) pooled ME over 10,000x to 220,000x dilution factors ($N$ = 4) and (B) pooled plasma over 3,000x to 30,000x ($N$ = 8). Data are the average of two replicates, with error bars representing the standard deviation. Average percent recovery for CA125 in (C) pooled ME samples ($N$ = 4) and (D) pooled plasma samples ($N$ = 8). Data are the average of two replicates, with error bars representing the standard deviation. Average percent recovery for CA125 in eight individual ME (E) and plasma (F) samples. Averages for low (2 pg/mL), medium (10 pg/mL), and high (100 pg/mL) CA125 spikes, with error bars representing the standard deviation across the three spike levels

#### S3.2 Comparing CA125 Levels between ME and Plasma Samples

We compared the CA125 levels in nine matched ME and plasma samples to assess the effect of centrifugation on measured CA125 levels (Supplemental Figure 4A; Supplemental Table 7). We performed spike-and-recovery experiments on each sample to validate measurement accuracy. Each sample was analyzed unspiked and after spiking with 2 pg/mL, 10 pg/mL, and 100 pg/mL recombinant CA125. We calculated percent recovery at each spike level and averaged across the three spike levels, using an acceptance criterion of 70-130%. Both ME and plasma were acceptable matrices for CA125 analysis. However, as anticipated, CA125 concentrations differed between the two matrices ($p = 0.01$) (Supplemental Figure 4A). The mean CA125 concentration in ME was 99,000 ± 63,000 pg/mL (5,500 ± 3,500 U/mL; see Supplemental Information, S4 for the conversion calculation), whereas the mean plasma CA125 concentration was 170,000 ± 137,000 pg/mL (9,500 ± 7,600 U/mL). Furthermore, the percentage change in CA125 concentration after centrifugation varied across individual samples, from 3% to 262%, with a median increase of 76% (Supplemental Figure 4B). The agreement between ME and plasma CA125 concentration was assessed using the Bland-Altman analysis (Supplemental Figure 4C). The mean systemic bias was -71,000 ± 112,000, with 95% limits of agreement ranging from -291,000 to 149,000; although CA125 concentration in ME was, on average, lower than in plasma, variability in the differences between the two biofluids was high. We hypothesize that the increase in CA125 levels in plasma after centrifugation results from volume reduction during centrifugation, which concentrates soluble CA125 in the plasma layer. Therefore, the plasma concentration becomes higher than in the more diluted, cell-containing, unprocessed ME matrix. These findings suggest that ME and plasma are not comparable matrices and should be validated separately.

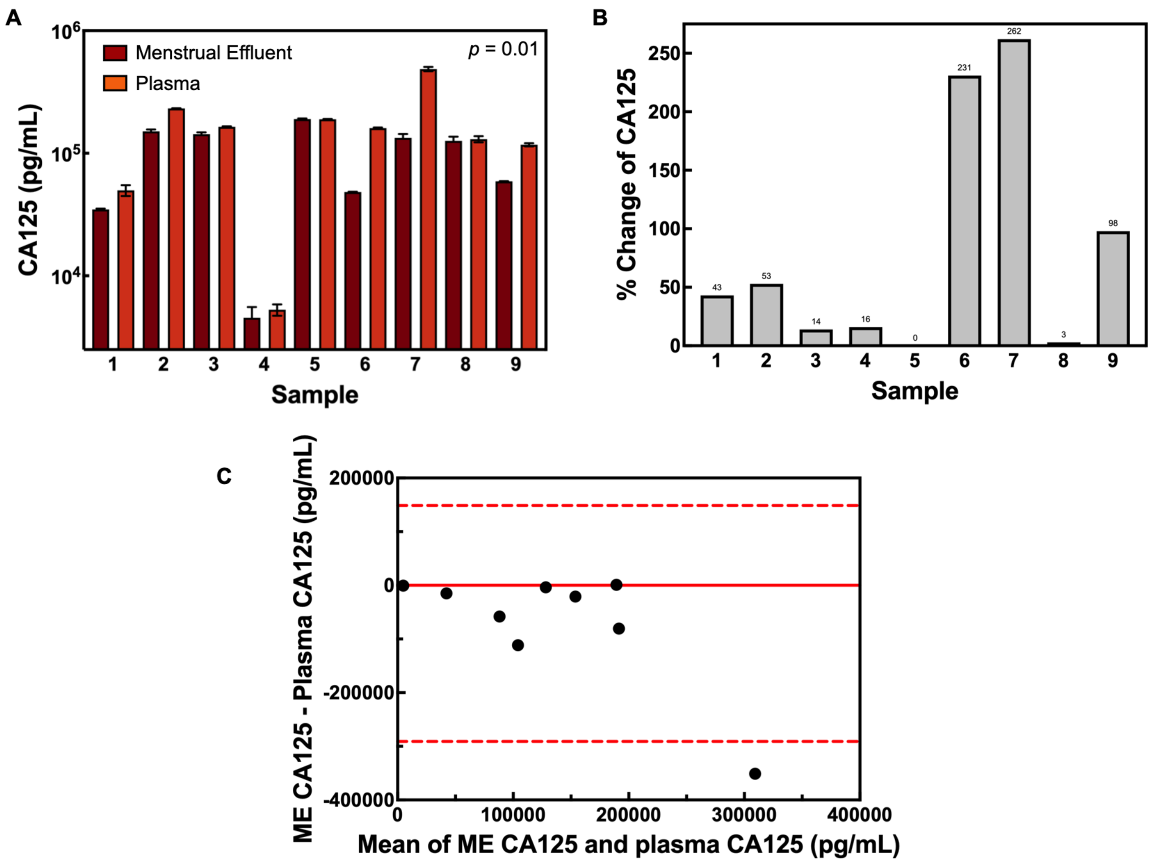

**Supplemental Figure 4:** (A) Comparison of CA125 levels in matched ME and plasma samples ($N = 9$ ME samples, $N = 9$ plasma samples). Each data point is an average of two replicates corrected by the dilution factor. Differences in medians CA125 concentration of ME and plasma samples were assessed by the Wilcoxon matched-pairs signed-rank test ($p < 0.05$) (B) Percentage change in CA125 concentration in ME and plasma samples. (C) Bland-Altman plot for CA125 levels measured in matched ME and plasma samples. The solid red line represents bias, and the dotted red lines indicate the 95% CI

**Supplemental Table 7:** Comparing CA125 concentration in nine matched ME and plasma samples

| Sample # | Sample Type | CA125 (pg/mL) | SD (pg/mL) | Adjusted CA125 (pg/mL) | Adjusted SD (pg/mL) | % Recovery for Sample Spiked with | | | |
|---|---|---|---|---|---|---|---|---|---|
| | | | | | | Low | Medium | High | Mean |
| 1 | ME | 0.580 | 0.010 | 34788 | 598 | 99% | 104% | 107% | 103% |
| | Plasma | 6.212 | 0.626 | 49699 | 5008 | 122% | 110% | 88% | 107% |
| 2 | ME | 2.521 | 0.087 | 151245 | 5217 | 141% | 120% | 104% | 122% |
| | Plasma | 28.990 | 0.033 | 231919 | 263 | 103% | 105% | 74% | 94% |
| 3 | ME | 2.390 | 0.080 | 143407 | 4786 | 89% | 96% | 92% | 92% |
| | Plasma | 20.523 | 0.140 | 164182 | 1121 | 81% | 85% | 108% | 91% |

| | | | | | | | | | |
|---|---|---|---|---|---|---|---|---|---|
| 4 | ME | 0.076 | 0.017 | 4555 | 999 | 100% | 99% | 107% | 102% |
| | Plasma | 0.659 | 0.071 | 5272 | 567 | 110% | 91% | 91% | 97% |
| 5 | ME | 3.163 | 0.040 | 189810 | 2382 | 85% | 88% | 94% | 89% |
| | Plasma | 23.617 | 0.123 | 188936 | 985 | 51% | 95% | 95% | 80% |
| 6 | ME | 0.805 | 0.005 | 48291 | 291 | 87% | 89% | 97% | 91% |
| | Plasma | 19.985 | 0.241 | 159877 | 1929 | 51% | 93% | 98% | 81% |
| 7 | ME | 2.229 | 0.164 | 133749 | 9870 | 85% | 92% | 96% | 91% |
| | Plasma | 60.601 | 2.506 | 484808 | 20045 | 155% | 89% | 90% | 111% |
| 8 | ME | 2.105 | 0.175 | 126285 | 10524 | 95% | 90% | 98% | 94% |
| | Plasma | 16.248 | 0.948 | 129983 | 7585 | 93% | 96% | 101% | 97% |
| 9 | ME | 0.985 | 0.003 | 59129 | 159 | 92% | 85% | 88% | 88% |
| | Plasma | 14.646 | 0.439 | 117170 | 3509 | 106% | 89% | 76% | 90% |

**Supplemental Table 8:** Aliquot-to-Aliquot Variability for ME ($N = 4$) and plasma ($N = 3$) samples

| Sample | Aliquot # | CA125 (pg/mL) | SD (pg/mL) | Adjusted CA125 (pg/mL) | Adjusted SD (pg/mL) | % Recovery for Sample Spiked with | | | |
|---|---|---|---|---|---|---|---|---|---|
| | | | | | | Low | Medium | High Spike | Mean |
| ME 1 | Aliquot 1 | 0.580 | 0.010 | 34788 | 598 | 99% | 104% | 107% | 103% |
| | Aliquot 2 | 0.325 | 0.006 | 19480 | 334 | 88% | 81% | 87% | 85% |
| ME 2 | Aliquot 1 | 2.390 | 0.080 | 143407 | 4786 | 89% | 96% | 92% | 92% |
| | Aliquot 2 | 1.509 | 0.134 | 90511 | 8024 | 90% | 85% | 91% | 89% |
| ME 3 | Aliquot 1 | 0.076 | 0.017 | 4555 | 999 | 100% | 99% | 107% | 102% |
| | Aliquot 2 | 0.036 | 0.016 | 2147 | 933 | 85% | 82% | 88% | 85% |
| ME 4 | Aliquot 1 | 13.767 | 1.082 | 826011 | 64891 | 122% | 87% | 90% | 100% |
| | Aliquot 2 | 13.869 | 0.290 | 832127 | 17423 | 143% | 99% | 93% | 112% |
| Plasma 1 | Aliquot 1 | 4.310 | 0.378 | 34479 | 3027 | 122% | 110% | 88% | 107% |
| | Aliquot 2 | 4.381 | 0.157 | 35052 | 1255 | 116% | 104% | 75% | 98% |
| Plasma 2 | Aliquot 1 | 11.112 | 0.251 | 88897 | 2010 | 120% | 94% | 87% | 100% |
| | Aliquot 2 | 11.249 | 1.534 | 89995 | 12269 | 132% | 87% | 94% | 104% |
| Plasma 3 | Aliquot 1 | 0.659 | 0.071 | 5272 | 567 | 110% | 91% | 91% | 97% |
| | Aliquot 2 | 0.665 | 0.037 | 5324 | 296 | 98% | 85% | 90% | 91% |

**Supplemental Table 9:** The effects of repeated freeze-thaw cycling (FT) on CA125 levels in plasma samples ($N = 8$)

| Plasma | FT # | CA125 (pg/mL) | SD (pg/mL) | Adjusted CA125 (pg/mL) | Adjusted SD (pg/mL) | % Recovery for Sample Spiked with | | | |
|---|---|---|---|---|---|---|---|---|---|
| | | | | | | Low | Medium | High Spike | Mean |
| 1 | 1 | 12.68 | 0.07 | 101456 | 584 | 134% | 86% | 95% | 105% |
| | 2 | 10.21 | 0.65 | 81651 | 5200 | 67% | 93% | 110% | 90% |
| | 3 | 11.36 | 1.25 | 90872 | 10017 | 113% | 99% | 92% | 101% |
| 2 | 1 | 19.98 | 0.24 | 159877 | 1929 | 51% | 93% | 98% | 81% |
| | 2 | 23.15 | 0.58 | 185210 | 4673 | 114% | 84% | 87% | 95% |
| | 3 | 19.11 | 0.82 | 152891 | 6577 | 118% | 87% | 95% | 100% |
| 3 | 1 | 14.95 | 1.16 | 119579 | 9242 | 97% | 94% | 103% | 98% |
| | 2 | 17.74 | 0.29 | 141933 | 2346 | 86% | 79% | 88% | 84% |
| | 3 | 16.51 | 0.50 | 132072 | 4038 | 57% | 81% | 95% | 78% |
| 4 | 1 | 6.56 | 0.09 | 52471 | 708 | 90% | 103% | 95% | 96% |
| | 2 | 15.77 | 0.45 | 126132 | 3619 | 85% | 85% | 113% | 94% |
| | 3 | 15.39 | 0.16 | 123158 | 1311 | 97% | 99% | 88% | 95% |
| 5 | 1 | 3.63 | 0.13 | 29022 | 1054 | 112% | 98% | 87% | 99% |
| | 2 | 3.90 | 0.04 | 31232 | 331 | 101% | 102% | 90% | 98% |
| | 3 | 3.73 | 0.11 | 29855 | 854 | 97% | 86% | 94% | 92% |
| 6 | 1 | 1.08 | 0.09 | 8670 | 721 | 106% | 106% | 113% | 108% |
| | 2 | 1.08 | 0.03 | 8679 | 201 | 98% | 85% | 85% | 89% |
| | 3 | 1.10 | 0.05 | 8774 | 420 | 87% | 94% | 93% | 91% |
| 7 | 1* | 6.21 | 0.63 | 49699 | 5008 | 122% | 110% | 88% | 107% |
| | 2 | 5.80 | 0.21 | 46396 | 1678 | 112% | 95% | 105% | 104% |
| | 3 | 6.20 | 0.24 | 49595 | 1931 | 110% | 107% | 104% | 107% |
| 8 | 1* | 28.99 | 0.03 | 231919 | 263 | 103% | 105% | 74% | 94% |
| | 2 | 28.23 | 1.36 | 225840 | 10904 | 188% | 84% | 96% | 123% |
| | 3 | 46.63 | 1.78 | 373006 | 14227 | 96% | 83% | 78% | 86% |

*(*) Indicates these results were obtained from a run with a high control, with a 21% error.*

**Supplemental Table 10:** Effect of storage conditions on sample stability

| Plasma | Storage Condition | CA125 (pg/mL) | SD (pg/mL) | Adjusted CA125 (pg/mL) | Adjusted SD (pg/mL) | % Recovery for Sample Spiked with | | | |
|---|---|---|---|---|---|---|---|---|---|
| | | | | | | Low | Medium | High Spike | Mean |
| 1 | 0H 20°C* | 185.175 | 6.458 | 1481397 | 51667 | 50% | 67% | 36% | 51% |
| | 6H 20°C* | 185.882 | 13.889 | 1487056 | 111111 | -43% | 54% | 43% | 18% |
| | 6h 4°C* | 163.144 | 7.826 | 1305148 | 62610 | 1138% | 269% | 57% | 488% |
| | 24h 4°C | 191.598 | 20.486 | 1532784 | 163886 | 72% | 144% | 53% | 90% |
| | 120h 4°C* | 194.522 | 7.490 | 1556179 | 59923 | -128% | -53% | 24% | -52% |
| 2 | 0H 20°C | 3.628 | 0.132 | 29022 | 1054 | 112% | 98% | 87% | 99% |
| | 6H 20°C | 3.333 | 0.195 | 26665 | 1557 | 105% | 94% | 103% | 101% |
| | 24H 20°C | 4.034 | 0.191 | 32269 | 1531 | 108% | 96% | 107% | 104% |
| | 120H 20°C | 3.509 | 0.060 | 28073 | 481 | 102% | 85% | 87% | 91% |
| | 6h 4°C | 3.206 | 0.247 | 25646 | 1974 | 128% | 104% | 115% | 116% |
| | 24h 4°C | 3.372 | 0.189 | 26975 | 1512 | 115% | 97% | 102% | 105% |
| | 120h 4°C | 3.592 | 0.007 | 28738 | 56 | 105% | 109% | 118% | 111% |
| 3 | 0H 20°C | 1.084 | 0.090 | 8670 | 721 | 106% | 107% | 113% | 109% |
| | 6H 20°C | 1.053 | 0.019 | 8423 | 153 | 103% | 104% | 121% | 109% |
| | 24H 20°C | 1.108 | 0.031 | 8861 | 249 | 102% | 99% | 110% | 104% |
| | 120H 20°C | 1.102 | 0.115 | 8815 | 917 | 95% | 100% | 91% | 95% |
| | 6h 4°C | 1.093 | 0.065 | 8741 | 520 | 97% | 94% | 106% | 99% |
| | 24h 4°C | 0.974 | 0.004 | 7796 | 35 | 87% | 96% | 88% | 90% |
| | 120h 4°C | 1.037 | 0.003 | 8297 | 26 | 99% | 96% | 103% | 99% |
| 4 | 6H 20°C | 35.895 | 0.277 | 287159 | 2217 | 73% | 62% | 88% | 74% |
| | 24H 20°C | 40.690 | 1.618 | 325519 | 12945 | 30% | 78% | 88% | 65% |
| | 6h 4°C | 33.899 | 2.326 | 271194 | 18609 | 61% | 82% | 91% | 78% |
| | 24h 4°C | 34.766 | 1.176 | 278130 | 9409 | 135% | 73% | 100% | 103% |
| | 120h 4°C | 35.363 | 1.079 | 282907 | 8628 | 179% | 90% | 93% | 121% |

*(*) Samples were excluded from further analysis as their percentage recovery fell outside the acceptance range of 70-130%.*

**Supplemental Table 11**: Longitudinal analysis of CA125 in ME Samples collected by one participant over three menstrual cycles.

| Month | Day of Cycle | CA125 (pg/mL) | SD (pg/mL) | Adjusted CA125 (pg/mL) | Adjusted SD (pg/mL) | % Recovery for Sample Spiked with | | | |
|---|---|---|---|---|---|---|---|---|---|
| | | | | | | Low | Medium | High | Mean |
| 1* | 2 | 191.6 | 20.5 | 1532784 | 163885 | 72% | 144% | 53% | 90% |
| 2 | 2 | 263.8 | 14.6 | 2110659 | 116738 | 177% | 183% | 19% | 127% |
| | 3 | 20.0 | 0.2 | 159877 | 1929 | 51% | 93% | 98% | 81% |
| | 4 | 14.9 | 1.2 | 119579 | 9242 | 97% | 94% | 103% | 98% |
| 3 | 3 | 60.6 | 2.5 | 484808 | 20045 | 155% | 89% | 90% | 111% |
| | 4 | 16.2 | 0.9 | 129983 | 7585 | 93% | 96% | 101% | 97% |

*(*) Sample analyzed after storage at 4°C for 24 hours.*

### S4. Conversion Calculation for CA125 Units (pg/mL to U/mL)

We analyzed CA125 concentrations using two calibrator reagents (assay 1: CA125 cat# 5609-05, R&D Systems; assay 2: Human CA125/MUC16 Quantikine ELISA kit, cat # DCA125**)** to provide an estimated relationship between the two measurement scales based on their limits of detection (LOD) in pg/mL and U/mL, respectively. The LOD was calculated as the concentration corresponding to three SD of the first calibrator added to the background signal. We averaged the ratio of CA125 concentrations from the two assays to estimate a conversion factor, which was then used to approximate CA125 concentrations in ME in U/mL (Supplemental Table 12).

**Supplemental Table 12:** Determining a conversion factor for pg/mL to U/mL using the CA125 concentration measured in

| LOD | Assay 1 (pg/mL) | Assay 2 (U/mL) | Ratio (U/mL per pg/mL) |
|---|---|---|---|
| 1 | 0.101 | 0.003 | 0.026 |
| 2 | 0.057 | 0.004 | 0.075 |
| 3 | 0.044 | 0.003 | 0.078 |
| 4 | 0.149 | 0.000 | 0.001 |
| 5 | 0.055 | 0.004 | 0.071 |
| 6 | 0.180 | 0.010 | 0.056 |
| 7 | 0.095 | 0.007 | 0.071 |
| 8 | 0.096 | 0.008 | 0.087 |
| 9 | 0.123 | 0.017 | 0.135 |
| 10 | 0.002 | 0.008 | 4.802 |
| 11 | 0.179 | 0.001 | 0.005 |
| 12 | 0.004 | 0.012 | 2.813 |
| 13 | 0.054 | 0.003 | 0.061 |
| 14 | 0.410 | 0.023 | 0.056 |
| 15 | 0.284 | 0.013 | 0.046 |
| Mean ratio (U/mL per pg/mL) | | | 0.559 |

### S5. Recommended Protocol for Menstrual Effluent Collection and Processing

This protocol is expected to provide a reproducible method for collecting and processing menstrual effluent and plasma, compatible with protein biomarker discovery.

#### Collection of Menstrual Effluent Samples

***Timing****:* ~3-5 hours

1. Provide participants with a self-sampling kit to collect menstrual effluent.
    a. A menstrual cup self-sampling kit should include at least one menstrual cup, a pair of disposable gloves, one blood collection tube, a biohazard bag, a menstrual cup protocol, a sampling information card, a post-collection survey and one ice pack.
    b. The Shordy menstrual cup is available in large and small sizes.
2. On the third day of the menstrual cycle, participants should wear a menstrual cup for 3-4 hours.
3. Place the ice pack in the freezer until the cup is removed.

*Note*: Participants can be asked to contact the laboratory during sample collection to provide an approximate drop-off time, allowing researchers time to prepare the instrument in advance.

4. After cup removal, participants should carefully transfer the menstrual effluent samples into the blood collection tube. Gently mix by inverting the collection tube 10 times.
5. Immediately place the blood collection tube in the biohazard bag with the ice pack for storage and transport. Samples should reach the laboratory within 8 hours of collection and be maintained at approximately 4 °C.
6. Record sample data in the sample information card (demographic information, collection day and time)

#### Menstrual Effluent Processing

***Timing***: ~ 40-70 minutes

*Note*: All handling of menstrual effluent should be under sterile conditions in a clean biosafety cabinet.

7. Upon arrival at the laboratory, begin sample processing as soon as possible after sample drop-off.

***Note*****:** Depending on the study objectives, select a biofluid and maintain it across all analyses as the matrix of interest. For example, menstrual effluent can be used for direct analysis or for extracting cellular or nucleic acid material. Plasma may be preferred for proteomic analysis, as removing cells and other debris improves sample clarity and enables higher reproducibility.

8. If delays occur, refrigerate at 4 °C to preserve sample stability and minimize changes to the protein biomarker concentration for up to 120 hours. Keep a detailed record of any delays that occur and the duration of the delay.

*Note*: A minimum of 10 µL is required for menstrual effluent analysis using Simoa technology. Samples < 10 µL should be regarded as insufficient for further processing.

9. Divide the sample into 100-250 µL aliquots. Record and label each aliquot.
    a. If interested in analyzing protein biomarkers in menstrual effluent, proceed to step 10. If interested in plasma, proceed to step 12.
10. After dividing the sample into aliquots, store them at −80°C as soon as possible for at least 24 hours prior to analysis. Avoid repeated freeze-thaw cycles.
11. Thaw the aliquot completely on ice for at least one hour before use.

#### Isolation of Plasma from Menstrual Effluent

***Timing***: ~ 40 minutes

12. Centrifuge at 2,000 × g at 4°C for 30 minutes
    a. If the sample fractionates, note the extent of hemolysis. If fractionation fails, the sample can be inverted 10 times and can be kept as ME
13. Carefully collect the plasma-like layer into 50 µL aliquots without disturbing the pellet. Plasma samples should also be stored at
14. Store the aliquots at −80°C promptly for at least 24 hours prior to analysis. Avoid repeated freeze-thaw cycles.
15. Thaw the aliquot completely on ice for at least one hour before use.

#### Record Keeping

***Timing***: ~10 minutes

16. Update the biobank log with the following information: participant identification number, collection day, menstrual cup wear time, collection tube type, drop-off time, sample volume, number of aliquots, pre-centrifugation delay duration and conditions, processing start time, and storage location and temperature.
